# Biomarker and therapeutic applications of a novel microRNA in early-stage knee osteoarthritis

**DOI:** 10.1101/2025.10.26.25338746

**Authors:** Madhu Baghel, Thomas G. Wilson, Trevor Banka, Jason Davis, Vasilios Moutzouros, Shabana Amanda Ali

## Abstract

Osteoarthritis is a debilitating joint disease with no known cure, underscoring the need for early detection and intervention. Given their potential disease-specificity, novel microRNAs are promising biomarkers and therapeutic targets. Novel microRNAs are identified from sequencing data using bioinformatics analyses. Here we aimed to characterize a novel microRNA (novel_miRNA_2; UCCCUGUUCGGGCGCCACU) in knee osteoarthritis. We show circulating novel_miRNA_2 accurately distinguishes early-stage knee osteoarthritis from both non-osteoarthritis and late-stage knee osteoarthritis. In knee osteoarthritis subchondral bone, novel_miRNA_2 functions as a canonical microRNA, downregulating target genes such as *RUNX2*. In male mice, novel_miRNA_2 mimic attenuates knee osteoarthritis when delivery begins in early stages, but not late stages. The decline in novel_miRNA_2 in late-stage knee osteoarthritis may be governed by Hedgehog signaling, which is known to upregulate genes such as *RUNX2* and exacerbate osteoarthritis. Our findings suggest endogenous novel_miRNA_2 can be used to detect early-stage knee osteoarthritis when exogenous delivery can mitigate disease.

## Introduction

Osteoarthritis (OA) is a debilitating joint disease currently impacting 7.6% of the global population^1^. Most commonly affecting the knee^1^, OA is characterized by phasic symptoms and progressive joint damage, including articular cartilage degradation, synovial inflammation, and subchondral bone remodeling^2^. There is no known cure for OA, and the molecular mechanisms that drive OA pathogenesis are incompletely understood, particularly in early-stage disease when prevention is still possible^3^. This points to a need for reliable biomarkers that can detect early-stage OA and for effective therapeutics that can mitigate disease progression^4^.

MicroRNAs are small non-coding RNAs with the potential to serve as both mechanistic biomarkers and therapeutic targets in OA^5^. Functioning to repress gene expression via direct binding to their target mRNAs^6^, microRNAs regulate key biological processes in joint tissues^7^. As biomarkers, circulating microRNAs are highly stable and can be linked to specific disease contexts^8^. As therapeutics, microRNAs can be modulated using small molecules to either enhance or inhibit their activity *in vivo* to mitigate disease processes^5,9^. Their dual function as mechanistic biomarkers and therapeutic targets can link diagnosis to treatment, making microRNAs strong candidates for precision medicine in OA.

Most studies to date have focused on established microRNAs that are cataloged in databases^10^ and characterized across different biological contexts^11^. In contrast, novel microRNAs are sequences that are newly discovered through bioinformatics analyses of microRNA-sequencing data^12^. Novel microRNAs align to the human reference genome, have a predicted hairpin secondary structure typical of microRNA precursors, and lack homology with established microRNAs in humans and other species^13^. With the potential to be completely unique to the biological context in which they are discovered, novel microRNAs are ideal for first-in-class microRNA biomarkers and therapeutics for OA. Despite their promise, the biological relevance and clinical utility of novel microRNAs in OA remains largely unknown.

In the first study to explore circulating novel microRNAs in OA, four candidates (novel_miRNA_1, novel_miRNA_2, novel_miRNA_3, and novel_miRNA_4) were found to be more prevalent in plasma from individuals with early-versus late-stage radiographic knee OA^14^. Here, we evaluate the biomarker potential of these four candidates in an independent patient cohort and prioritize novel_miRNA_2. Using both primary human knee OA tissues and a surgical mouse model of knee OA, we define expression profiles and therapeutic effects of novel_miRNA_2. Together, our findings suggest that novel_miRNA_2 is a promising biomarker for early-stage knee OA that functions as a canonical microRNA to prevent disease by repressing aberrant bone-related processes.

## Results

### Circulating novel_miRNA_2 distinguishes early-stage knee OA

Four novel microRNAs were previously reported in the plasma of individuals with early-versus late-stage radiographic knee OA^14^ (**Figure 1A**). To validate these findings, we leveraged the Henry Ford Health (HFH) OA Cohort, a repository of human biofluids, tissues, and clinicodemographic data from individuals with symptomatic knee OA undergoing arthroscopy or arthroplasty. Participants were stratified by radiographic severity using the Kellgren-Lawrence (KL) grading system, with KL grade 0 or 1 defined as early-stage knee OA^15^ and KL grade 3 or 4 as late-stage knee OA, consistent with previous definitions^14^. Non-OA plasma samples were obtained from the “Control Cohort” of the Osteoarthritis Initiative (OAI)^16^ and included individuals with no symptomatic or radiographic evidence of knee OA.

**Figure 1.**
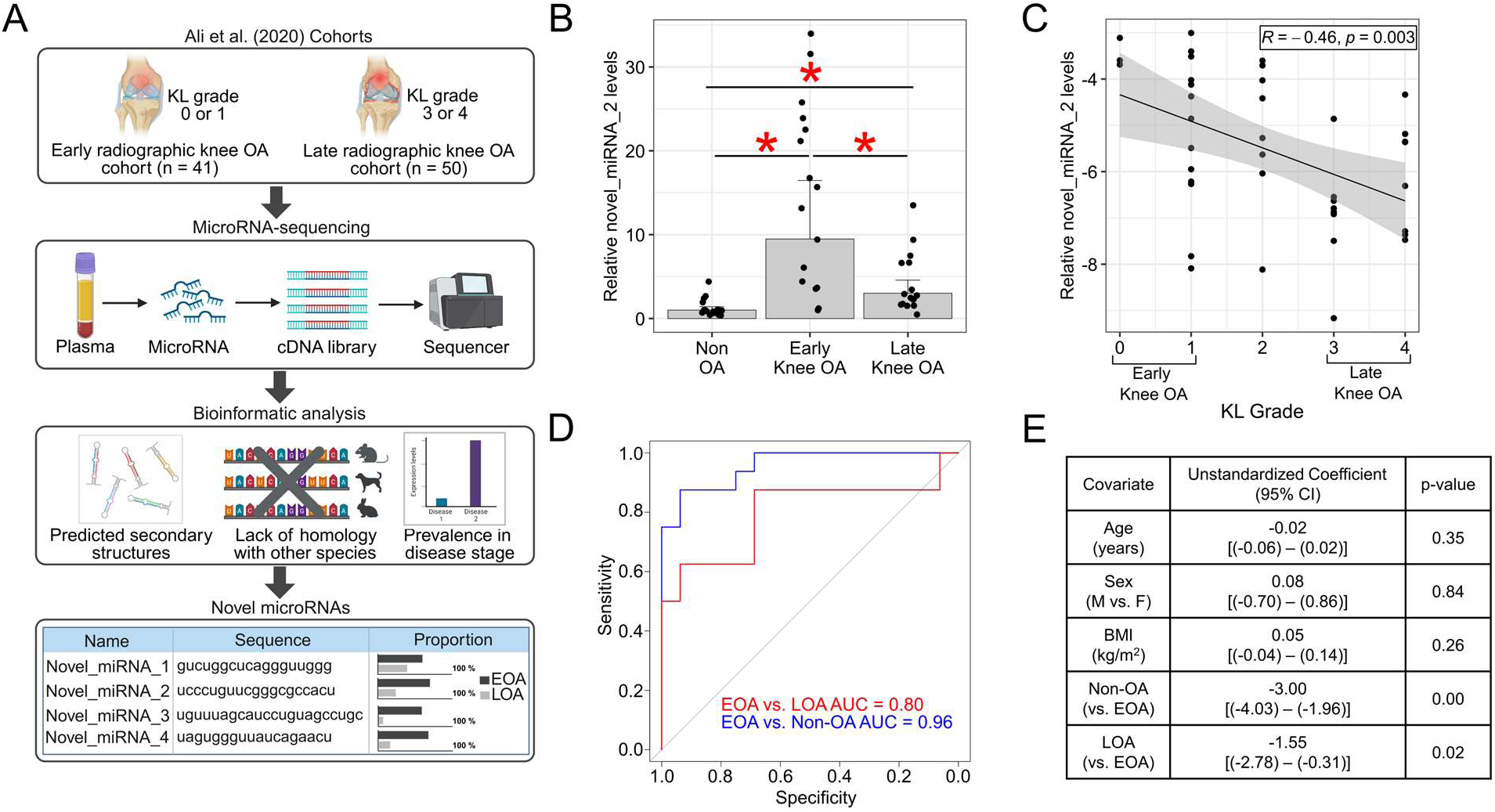
Circulating novel_miRNA_2 distinguishes early-stage knee OA. A) Overview of the approach used by Ali et al. (2020)^14^ to identify four novel microRNAs prevalent in early-stage knee OA plasma. Image created in BioRender. B) Relative plasma levels of novel_miRNA_2 in the HFH OA Cohort. Values represent the fold-change relative to non-OA controls (n = 16/group). Bars represent mean ± 95% confidence interval. *p < 0.05 as determined by one-way ANOVA with Tukey’s HSD post hoc test. C) Pearson correlation of plasma novel_miRNA_2 levels versus Kellgren-Lawrence (KL) grade (n = 40). Data fitted from novel_miRNA_2 plasma levels relative to a reference microRNA (miR-24-3p). Black line = fitted linear model. Grey shaded region = 95% confidence interval. D) Area under the receiver operating characteristic curve (AUC) analysis evaluating the accuracy of novel_miRNA_2 plasma levels in distinguishing early-stage knee OA (EOA) from non-OA controls (blue line) and late-stage knee OA (LOA; red line). E) Multiple linear regression analysis evaluating the association of covariates with plasma novel_miRNA_2 levels. M = male, F = female, BMI = body mass index, 95% CI = confidence interval. Statistical significance for regression coefficients determined by two-sided t-tests. P-values: B) p = 0.000 (Early Knee OA vs. Non-OA), p = 0.002 (Early Knee OA vs. Late Knee OA), p = 0.003 (Late Knee OA vs. Non-OA).

Performing real-time PCR (RT-qPCR) using custom-designed TaqMan assays, no significant differences were observed for novel_miRNA_1, novel_miRNA_3, or novel_miRNA_4 (**Supplemental Figure 1**), but novel_miRNA_2 was significantly elevated in early-stage knee OA plasma compared to non-OA controls and late-stage knee OA (**Figure 1B**). When including KL grade 2 samples, plasma levels of novel_miRNA_2 showed a significant inverse correlation with KL grade (**Figure 1C**). Based on these findings, we evaluated the potential of novel_miRNA_2 to distinguish early-stage knee OA using area under the receiver operating characteristic curve analysis (AUC). Novel_miRNA_2 showed excellent accuracy in discriminating early-stage knee OA from non-OA and good accuracy from late-stage knee OA (**Figure 1D**), as well as good accuracy in distinguishing late-stage knee OA from non-OA (**Supplemental Figure 2**). Using multivariable linear regression, we confirmed there were no associations between novel_miRNA_2 plasma levels and age, sex, or body mass index (BMI), but significant associations with early-stage knee OA (**Figure 1E**). Collectively, these findings prioritize novel_miRNA_2 as a promising circulating biomarker for early-stage knee OA.

### Novel_miRNA_2 functions as a canonical microRNA

Since novel_miRNA_2 was predicted to be a microRNA using only bioinformatics analyses^14^, we aimed to experimentally verify its authenticity based on known properties of canonical microRNAs^6^. MicroRNA biogenesis begins with transcription of a primary (“pri-”) transcript from a host gene, which folds into a hairpin structure^6^. This precursor undergoes enzymatic cleavages to generate mature microRNAs that are loaded into Argonaute 2 (AGO2)-containing RNA-induced silencing complexes (RISCs) to mediate repression of target genes^6^ (**Figure 2A**). Prior to experimentation, we aligned the predicted novel_miRNA_2 precursor to the human reference genome (GRCh38) and identified its host gene as Zinc Finger Protein 773 (*ZNF773*), with novel_miRNA_2 located on the reverse strand (**Figure 2B**). We also identified the mature novel_miRNA_2 sequence on the 5’ arm of the precursor molecule, which designates it as a “-5p” microRNA by standard conventions^17^ (**Figure 2B**).

**Figure 2.**
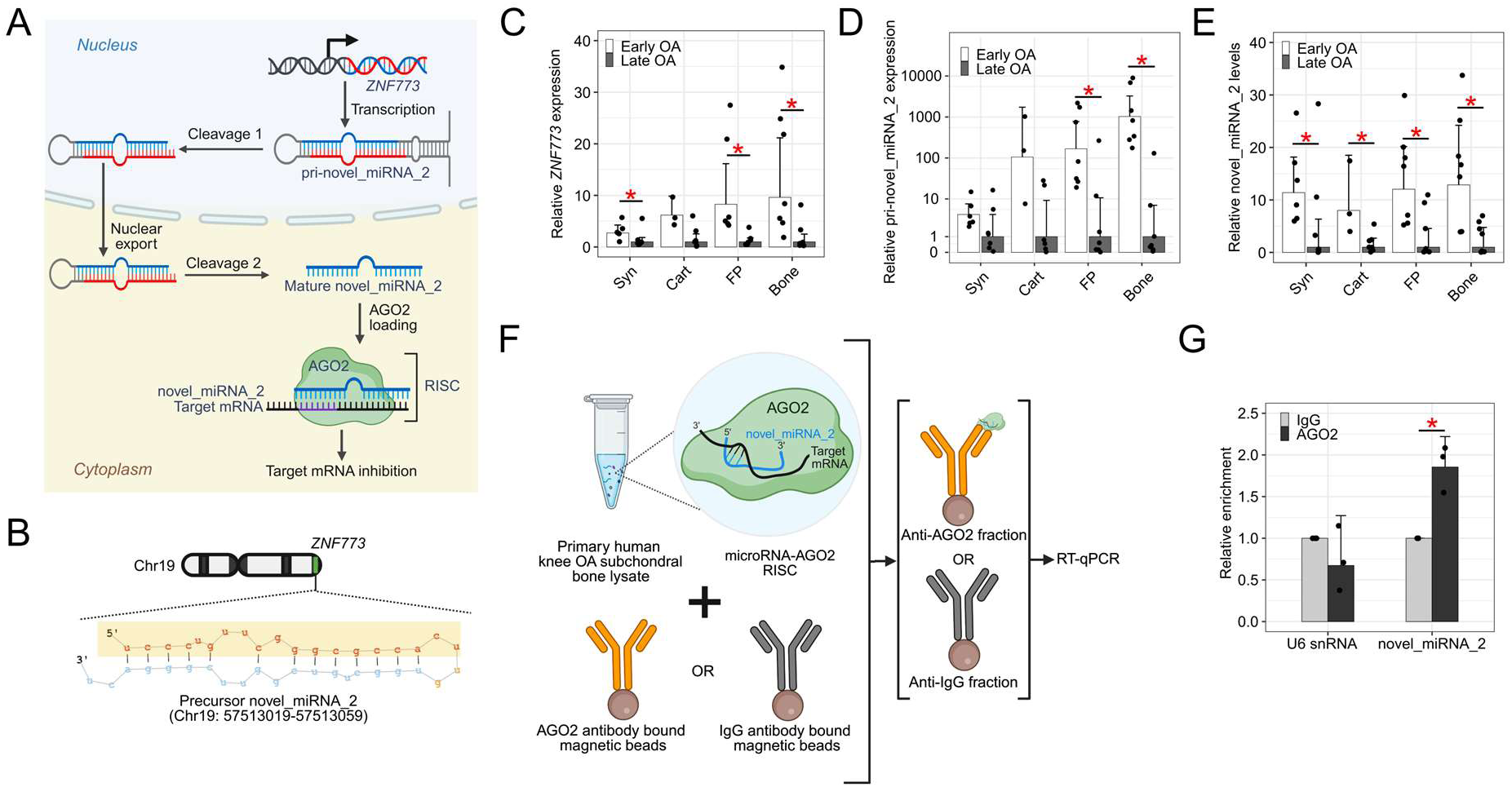
Novel_miRNA_2 functions as a canonical microRNA. A) Overview of the putative biogenesis of novel_miRNA_2. RISC = RNA-induced silencing complex. Image created in BioRender. B) Predicted secondary structure of the novel_miRNA_2 precursor located in the *ZNF773* host gene on human chromosome 19 (Chr19). Mature novel_miRNA_2 (highlighted yellow) originates from the 5’ arm of the precursor. Image created in BioRender. C) *ZNF773*, D) pri-novel_miRNA_2, and E) mature novel_miRNA_2 levels in early-(n = 3-7) and late-stage (n = 7) human knee OA tissues. Data represent fold-change values relative to the average levels in corresponding late-stage knee OA tissues. Bars represent mean ± 95% confidence interval. *p < 0.05 as determined by two-sided Mann-Whitney U-test (*ZNF773*) or two-sided Welch’s T-test (pri-novel_miRNA_2 and novel_miRNA_2). “Bone” = subchondral bone, “FP” = infrapatellar fat pad, “Cart” = articular cartilage, “Syn” = synovium. F) Overview of AGO2-RNA immunoprecipitation assay in knee OA subchondral bone. Image created in BioRender. G) Relative enrichment of novel_miRNA_2 and U6 small nuclear RNA (snRNA) negative control in IgG-or AGO2-bound fractions as measured by RT-qPCR. Data represent fold-change values relative to the corresponding IgG-bound fraction (n = 3). Bars represent mean ± 95% confidence. *p < 0.05 as determined by two-sided Welch’s T-test. P-values: C) p = 0.007 (Bone), p = 0.001 (FP), p = 0.067 (Cart), p = 0.035 (Syn). D) p = 0.000 (Bone), p = 0.005 (FP), p = 0.055 (Cart), p = 0.109 (Syn). E) p = 0.018 (Bone), p = 0.018 (FP), p = 0.017 (Cart), p = 0.042 (Syn). G) p = 0.021 (novel_miRNA_2), p = 0.347 (U6 snRNA).

We leveraged the steps of microRNA biogenesis to identify the human knee OA tissues that express novel_miRNA_2. Specifically, we quantified *ZNF773*, pri-novel_miRNA_2, and mature novel_miRNA_2 in subchondral bone, infrapatellar fat pad (“fat pad”), articular cartilage, and synovium using RT-qPCR. In early-versus late-stage knee OA tissues, *ZNF773* was significantly higher in subchondral bone, fat pad, and synovium (**Figure 2C**), pri-novel_miRNA_2 was higher in subchondral bone and fat pad (**Figure 2D**), and mature novel_miRNA_2 was higher in subchondral bone, fat pad, synovium, and articular cartilage (**Figure 2E**). Since the mature form is derived from the primary transcript, we performed correlation analyses between mature novel_miRNA_2 and pri-novel_miRNA_2 levels and found significant positive associations in subchondral bone (n = 14, R = 0.69, p = 0.01) and fat pad (n = 14, R = 0.67, p = 0.01). To assess incorporation of novel_miRNA_2 into RISCs, we performed an AGO2-RNA immunoprecipitation assay in human knee OA subchondral bone (**Figure 2F**). We found significant enrichment of novel_miRNA_2 in AGO2-bound fractions compared with IgG fractions, while there was no enrichment of negative control U6 small nuclear RNA (**Figure 2G**). Taken together, these results show that novel_miRNA_2 is elevated in early-versus late-stage knee OA tissues and follows canonical microRNA biogenesis most consistently in subchondral bone.

### Novel_miRNA_2 inhibits target genes in knee OA subchondral bone

To elucidate molecular mechanisms of novel_miRNA_2 in knee OA, we sought to identify its direct target genes. MicroRNAs inhibit gene expression through complementary seed-sequence binding, most often within the 3’ untranslated region (UTR) of a target mRNA^6^. Using an *in silico* approach^18^, we first identified 2,951 human genes that contained at least one putative 7mer or 8mer novel_miRNA_2 seed-sequence binding site in their 3’ UTR (**Figure 3A**). Second, we evaluated binding site stability by calculating the Gibbs free energy (ΔG)^19^ for the 70-nucleotide flanking regions of each binding site, comparing them with 3’ UTR-wide averages. Sites with greater than average flanking ΔG were retained, indicative of less stable mRNA secondary structures and higher accessibility for RISC binding^19^, resulting in 1,463 genes (**Figure 3A**). Third, to focus on OA, we cross-referenced this list with bone-related datasets curated in OsteoDIP^20^, leaving 87 genes (**Figure 3A**). Finally, we performed pathway analysis^21^ on this subset and found enrichment of curated ‘Diseases and Functions’ and ‘Canonical Pathways’ that relate to OA (**Supplemental Figure 3**), resulting in a final shortlist of 13 candidate target genes (**Figure 3B**).

**Figure 3.**
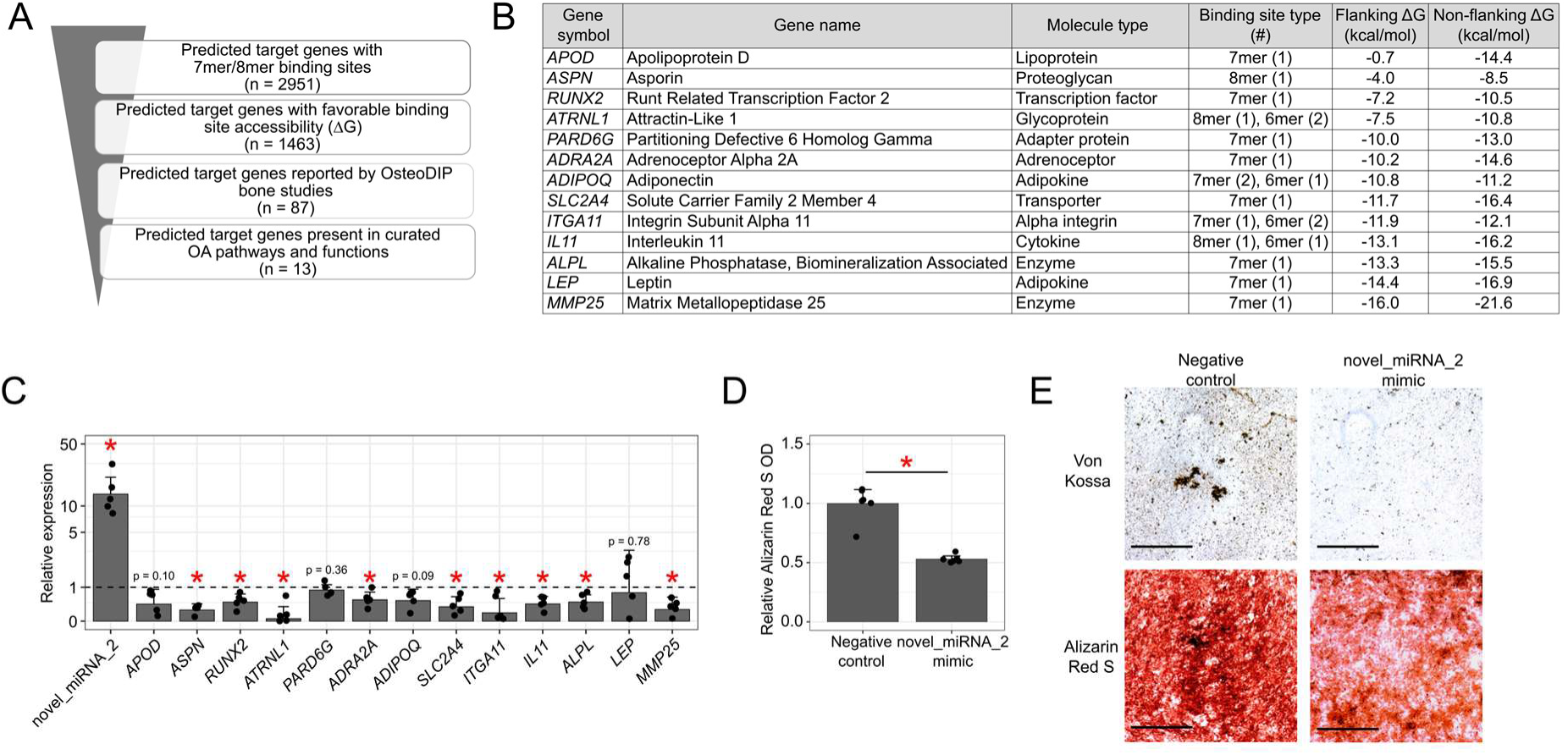
Novel_miRNA_2 inhibits target genes in knee OA subchondral bone. A) Strategy to prioritize predicted novel_miRNA_2 target genes in OA. ΔG = Gibbs free energy. B) List of the 13 shortlisted target genes with features of their predicted novel_miRNA_2 binding sites, ranked by predicted flanking region ΔG. C) RT-qPCR in knee OA subchondral bone transfected with novel_miRNA_2 mimic or negative control (n = 5). Data represented relative to negative control-treated tissues (dashed line). Bars represent mean ± 95% confidence interval. *p < 0.05 versus matched negative control. D) Spectrophotometric quantification of Alizarin Red S staining (n = 6 technical replicates). Data represented relative to negative control-treated cells. Bars represent mean ± 95% confidence interval. *p < 0.05. E) Representative images of MC3T3-E1 cells stained with von Kossa or Alizarin Red S dyes to visualize mineralization following transfection with novel_miRNA_2 mimic or negative control. Scale bars = 1 mm. All significance assessed by paired two-sided Student’s T-test. P-values: C) p = 0.000 (novel_miRNA_2), p = 0.007 (*ASPN*), p = 0.031 (*RUNX2*), p = 0.040 (*ATRNL1*), p = 0.041 (*ADRA2A*), p = 0.031 (*SLC2A4*), p = 0.047 (*ITGA11*), p = 0.019 (*IL11*), p = 0.023 (*ALPL*), p = 0.041 (*MMP25*). D) p = 0.000.

To validate these predicted targets, we transfected human knee OA subchondral bone explants with a custom-designed novel_miRNA_2 mimic to enhance its levels, then assessed expression of the 13 genes (**Figure 3C**). Nine genes (*ASPN*, *RUNX2*, *ATRNL1*, *ADRA2A*, *SLC2A4*, *ITGA11*, *IL11*, *ALPL*, and *MMP25*) were significantly downregulated with novel_miRNA_2 mimic treatment versus negative control treatment, and four genes (*APOD*, *PARD6G*, *ADIPOQ,* and *LEP*) were not significantly different (**Figure 3C**). These results are consistent with the expected inhibitory effect of microRNAs on their direct target genes^6^. To assess functional effects of novel_miRNA_2 in subchondral bone, we explored bone mineralization, a process required for the osteophyte formation and subchondral sclerosis observed in OA^22^. In murine pre-osteoblast cells transfected with novel_miRNA_2 mimic, there was a significant reduction in mineral deposition compared to negative control (**Figure 3D, E**). Together, these results suggest that novel_miRNA_2 functions to regulate target genes and bone-related processes such as mineralization in knee OA subchondral bone.

### Novel_miRNA_2 has a protective effect in early-stage knee OA

To assess the effect of novel_miRNA_2 on knee OA *in vivo*, we leveraged the partial medial meniscectomy (PMX) mouse model^23^. Since species specificity was a criterion used to discover novel_miRNA_2, it is predicted to occur in humans but not mice^13,14^. Consistent with this, we found no matching sites upon aligning either the precursor or mature novel_miRNA_2 sequences to the mouse reference genome (GRCm39). We were therefore able to precisely control novel_miRNA_2 levels in mice using mimic treatments. Based on our data showing novel_miRNA_2 is elevated in circulation during early-stage knee OA (**Figure 1B**), and previous literature characterizing OA pathology in PMX mice over time^24^, we compared intravenous treatment beginning at two different timepoints (**Figure 4A**). Early treatment began one week post-operatively, when cartilage degradation, osteophyte formation, and synovitis are all underway, while late treatment began four weeks post-operatively, when these processes are more established^24^ (**Figure 4A**). To first confirm that the novel_miRNA_2 mimic was reaching knee tissues following intravenous delivery, we performed *in situ* hybridization and observed a positive signal in the subchondral bone of mimic-treated mice one week post-surgery compared to negative control treatment (**Figure 4B**). Notably, signal intensity was greater in PMX knees compared to sham knees (**Figure 4B**), suggesting that circulating novel_miRNA_2 accumulates in mouse tissues during knee OA, as observed in human tissues (**Figure 2E**).

**Figure 4.**
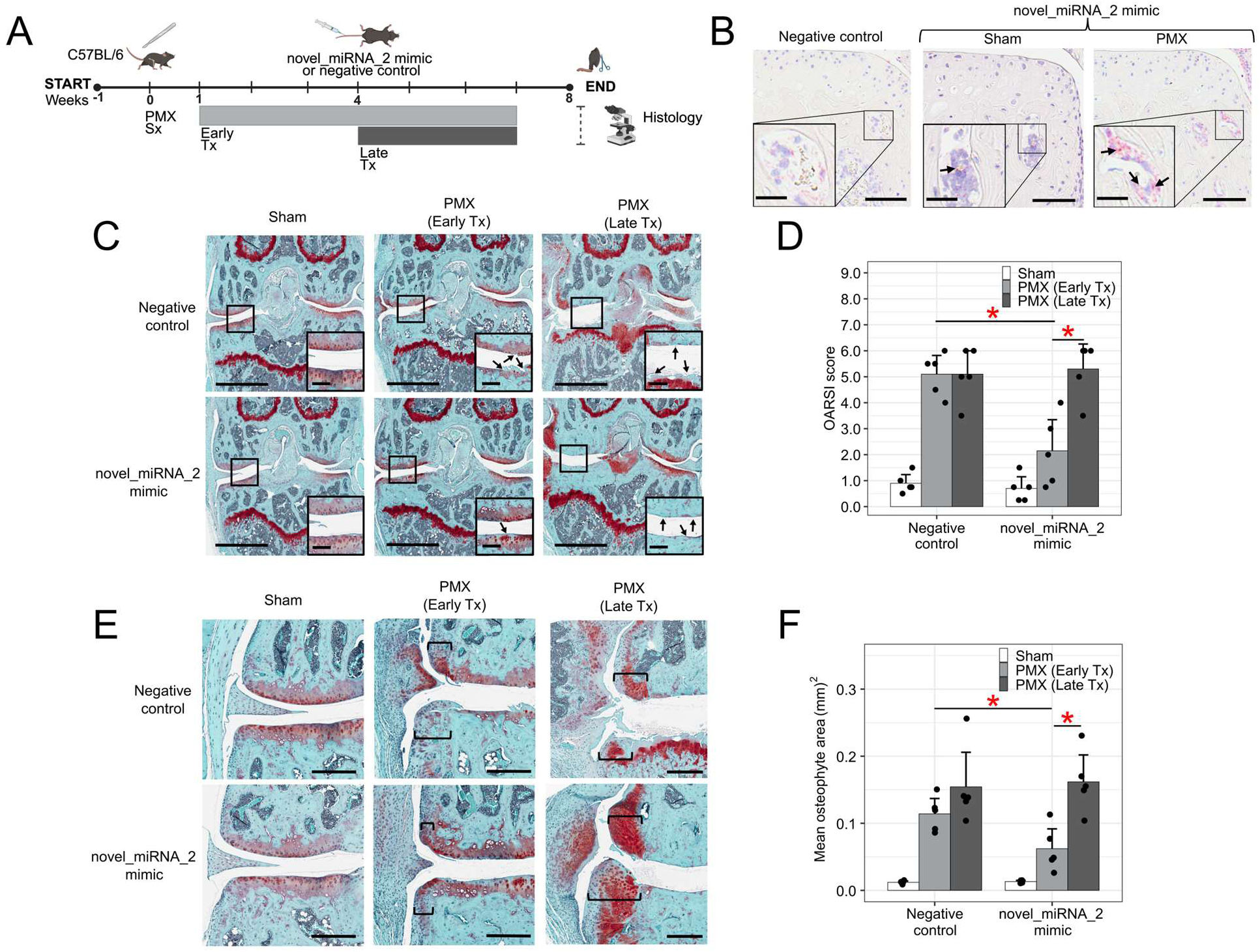
Novel_miRNA_2 has a protective effect in early-stage knee OA. A) Overview of the experimental design for mouse surgeries and novel_miRNA_2 mimic treatments. PMX = partial medial meniscectomy. Sx = surgery. Tx = treatment. Image created in BioRender. B) Representative images from *in situ* hybridization depicting localization of novel_miRNA_2 mimic in mouse knee subchondral bone. Scale bars = 200 µm. Insets show magnified regions (scale bars = 50 µm, arrows = positive signal). C) Representative coronal sections of mouse operated knee, stained with Safranin-O. Scale bars = 1 mm. Insets show magnified regions (scale bars = 100 µm, arrows = examples of cartilage damage). D) OARSI scoring of PMX and sham mice following novel_miRNA_2 mimic or control treatments (n = 5). Bars represent mean maximal quadrant score ± 95% confidence interval. *p < 0.05. E) Representative coronal sections of mouse operated knee demonstrating osteophyte burden. Scale bars = 250 µm. Brackets = approximate osteophyte diameter. F) Osteophyte area in PMX and sham mice following novel_miRNA_2 mimic or control treatments (n = 5). Datapoints represent the mean of the medial tibial and medial femoral osteophyte area for each mouse. Bars represent the mean osteophyte area ± 95% confidence interval, *p < 0.05. All significance assessed by two-sided Mann-Whitney U-test with Benjamini-Hochberg multiple testing correction. P-values: D) p = 0.030 [novel_miRNA_2 mimic: PMX (Early Tx) vs. PMX (Late Tx)], p = 0.016 [PMX (Early Tx): Negative control vs. novel_miRNA_2 mimic]. F) p = 0.024 [novel_miRNA_2 mimic: PMX (Early Tx) vs. PMX (Late Tx)], p = 0.032 [PMX (Early Tx): Negative control vs. novel_miRNA_2 mimic].

To assess knee OA severity, we evaluated cartilage degradation, osteophyte burden, and synovitis as well-characterized features in the PMX model^24^. Compared to negative control treatment, we found novel_miRNA_2 mimic reduced cartilage degradation (**Figure 4C, D**) and osteophyte burden (**Figure 4E, F**) in PMX knees, but only when treatments began at earlier stages and not later stages. No significant effect on synovitis was observed with either treatment strategy (**Supplemental Figure 4**). These findings suggest novel_miRNA_2 likely functions to mitigate tissue-specific disease processes in early stages of OA, but cannot reverse those processes once established in later stages.

### Hedgehog signaling may regulate novel_miRNA_2 levels in knee OA

Given the disease stage-dependent biomarker (**Figure 1**) and therapeutic (**Figure 4**) patterns exhibited by novel_miRNA_2, we sought to identify factors governing its levels in early-versus late-stage knee OA. Taking an *in silico* approach to identifying transcription factor binding sites within the promoter region of the *ZNF773* host gene, we found three GLI consensus binding motifs located within 2.5 kb of novel_miRNA_2 (**Figure 5A**). GLI transcription factors are the primary effectors of the Hedgehog (Hh) signaling pathway, which is known to exacerbate knee OA^25^. Since Hh signaling upregulates Runt Related Transcription Factor 2 (*RUNX2)*^25^, a regulator of bone formation^26^ and a predicted target gene of novel_miRNA_2 (**Figure 3B, C**), we hypothesized that Hh signaling governs novel_miRNA_2 levels in knee OA subchondral bone.

**Figure 5.**
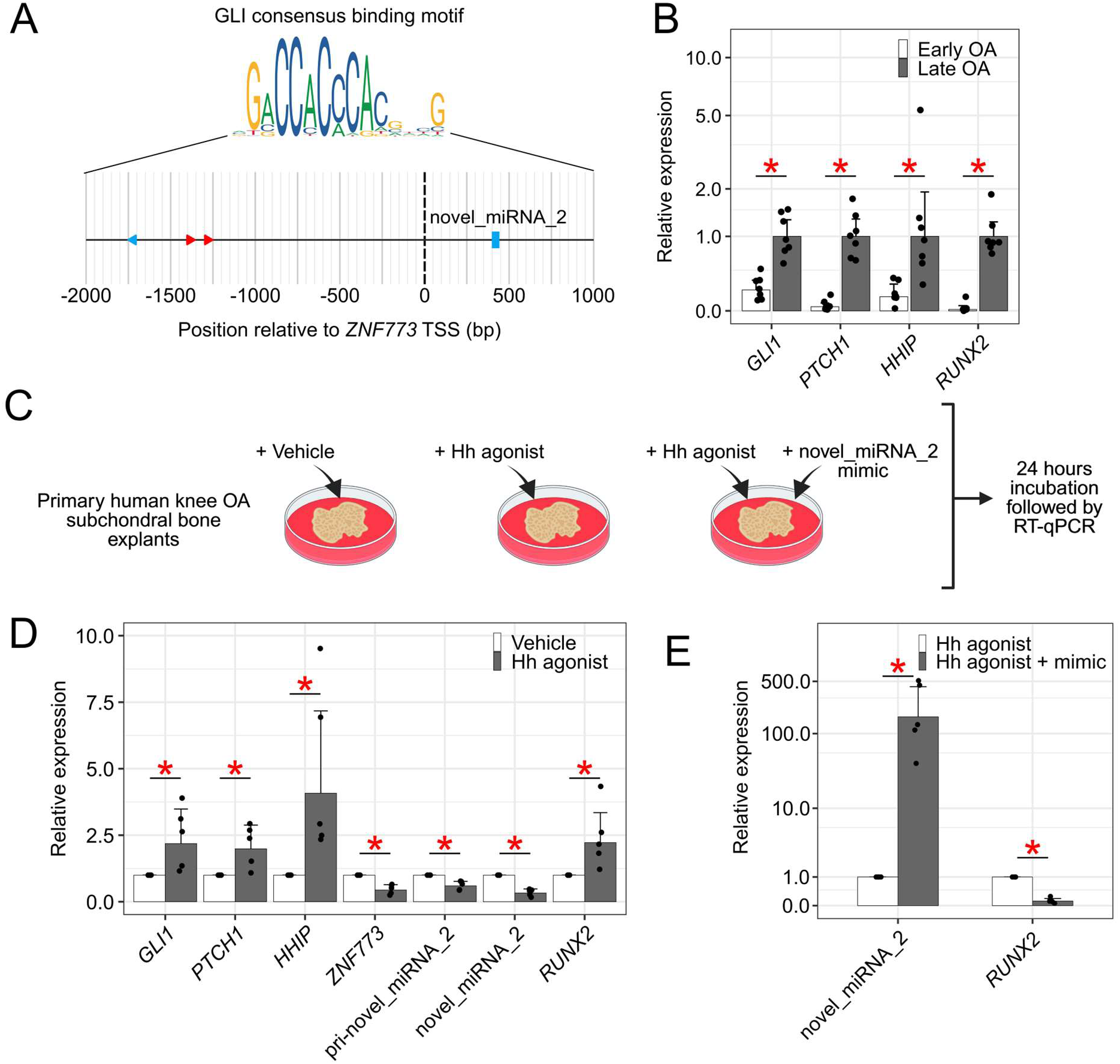
Hedgehog (Hh) signaling may regulate novel_miRNA_2 levels in knee OA. A) GLI consensus binding motifs in the region 2.5 kb upstream of novel_miRNA_2 within *ZNF773*. Red = positive strand, blue = negative strand. TSS = transcription start site. B) RT-qPCR in human knee OA subchondral bone (n = 7/group). Data represent fold-change relative to late-stage knee OA tissues. Bars represent mean ± 95% confidence interval. *p < 0.05 as determined by two-sided Welch’s T-test. C) Experimental design for Hh agonist (purmorphamine) treatments. Vehicle = dimethyl sulfoxide (DMSO). Image created in BioRender. D) RT-qPCR in late-stage knee OA subchondral bone explants following Hh agonist treatment (n = 5/group). Data represent fold-change relative to paired vehicle (DMSO) control. Bars represent mean ± 95% confidence interval. *p < 0.05. E) RT-qPCR in late-stage knee OA subchondral bone explants (n = 5/group) following treatment with Hh agonist and concurrent novel_miRNA_2 mimic (“mimic”). Data represent fold-change relative to Hh agonist alone. Bars represent mean ± 95% confidence interval. *p < 0.05 as determined by two-sided paired Student’s T-test. P-values: B) p = 0.000 (*GLI1*), p = 0.000 (*PTCH1*), p = 0.003 (*HHIP*), p = 0.001 (*RUNX2*). D) p = 0.030 (*GLI1*), p = 0.022 (*PTCH1*), p = 0.008 (*HHIP*), p = 0.013 (*ZNF773*), p = 0.015 (pri-novel_miRNA_2), p = 0.005 (novel_miRNA_2), p = 0.019 (*RUNX2*). E) p = 0.000 (novel_miRNA_2), p = 0.001 (*RUNX2*).

To investigate Hh signaling in early-versus late-stage human knee OA subchondral bone, we measured the Hh target genes GLI Family Zinc Finger 1 (*GLI1*), Patched 1 (*PTCH1*), and Hedgehog Interacting Protein (*HHIP*) as markers of pathway activity^25,27^. All three targets were lower in early-versus late-stage knee OA subchondral bone, as were levels of *RUNX2* (**Figure 5B**), consistent with the positive relationship previously reported between Hh signaling and *RUNX2* levels^25^. Together with the higher levels of *ZNF773*, pri-novel_miRNA_2, and mature novel_miRNA_2 we observed in early-stage knee OA subchondral bone (**Figure 2C-E**), these findings point to an inverse relationship between Hh signaling and novel_miRNA_2.

To assess the extent to which Hh signaling regulates novel_miRNA_2 levels, we treated human knee OA subchondral bone explants with the Hh agonist purmorphamine^28^ or vehicle control (**Figure 5C**). Hh pathway activation by purmorphamine was confirmed by an increase in *GLI1*, *PTCH1*, and *HHIP* compared to vehicle control (**Figure 5D**). Concurrently, we observed significant reductions in *ZNF773*, pri-novel_miRNA_2, and mature novel_miRNA_2, along with increased expression of *RUNX2* (**Figure 5D**). These findings suggest that Hh signaling can reduce novel_miRNA_2 and remove the inhibitory effect novel_miRNA_2 has on target genes such as *RUNX2.* Consistent with this, when novel_miRNA_2 levels were restored via mimic treatment (**Figure 5C**), *RUNX2* expression decreased, even in the presence of Hh agonist (**Figure 5E**). Taken together, these findings suggest that Hh signaling may govern the decline in novel_miRNA_2 levels from early-to late-stage knee OA, and when restored, novel_miRNA_2 can mitigate the effect of Hh signaling on *RUNX2* in knee OA subchondral bone (**Figure 6**).

**Figure 6.**
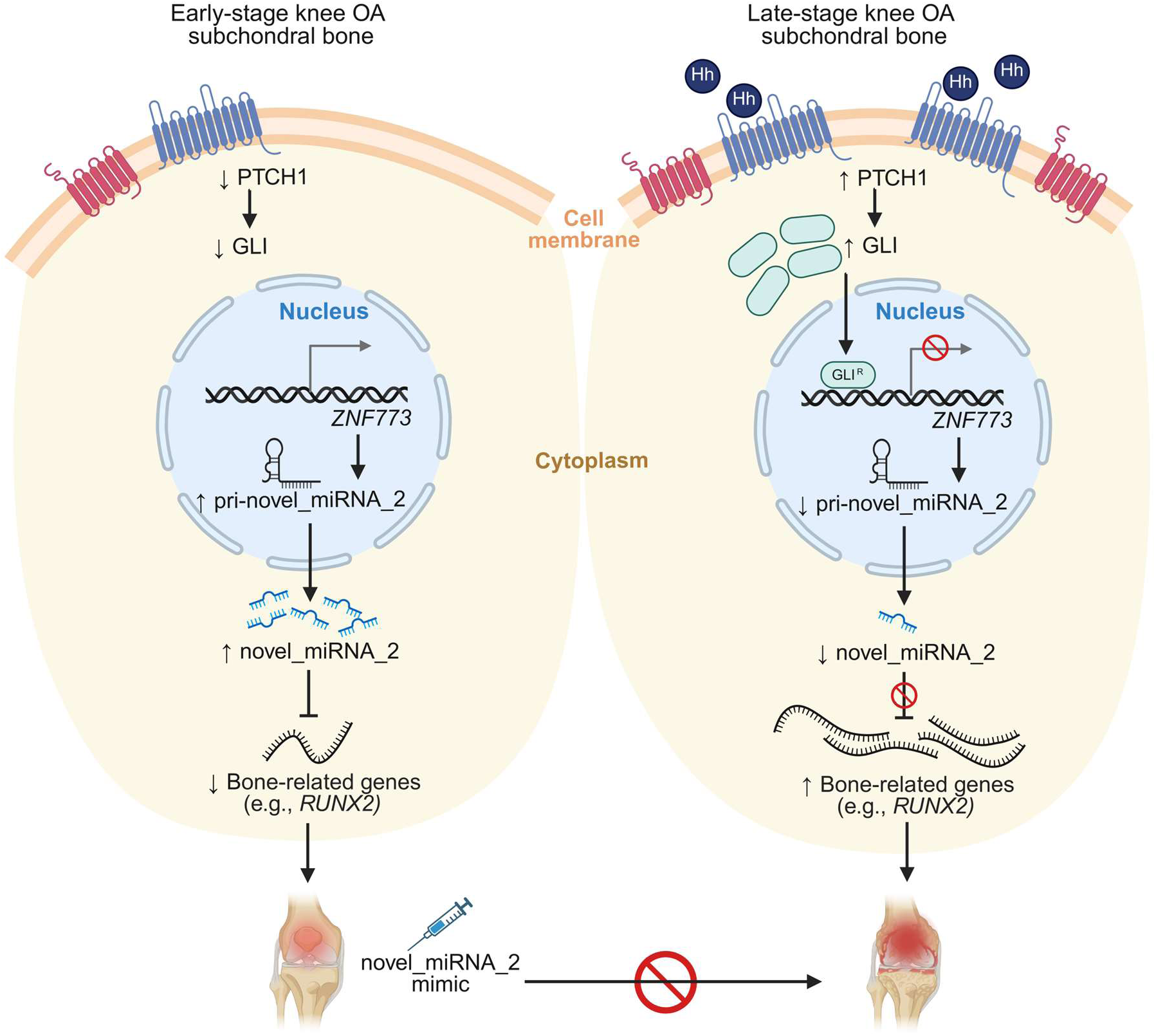
Novel_miRNA_2 may mediate the effects of Hedgehog (Hh) signaling on *RUNX2* in knee OA subchondral bone. In early-stage knee OA subchondral bone (left), reduced Hh pathway activity coincides with elevated novel_miRNA_2 biogenesis and expression levels, which can repress target genes such as *RUNX2*. In late-stage knee OA subchondral bone (right), increased Hh pathway activity may suppress novel_miRNA_2 levels, potentially via GLI repressor isoforms, and allow *RUNX2* to be elevated. When novel_miRNA_2 mimic is delivered beginning in early stages of OA, its protective effects are maintained (bottom). Image created in BioRender.

## Discussion

Here we characterize a novel microRNA with the potential to be unique to OA. The novel_miRNA_2 sequence has been reported by two studies profiling circulating novel microRNAs in OA^14,29^, but has not been reported in other studies profiling novel microRNAs in non-OA human biospecimens^12,30,31^. We show circulating novel_miRNA_2 can distinguish early-stage knee OA from both non-OA controls and late-stage knee OA, suggesting utility as a minimally invasive biomarker. In human knee OA subchondral bone, novel_miRNA_2 functions as a canonical microRNA that is transcribed from the *ZNF773* host gene, processed from primary transcript into mature form, and incorporated into AGO2-containing RISCs to inhibit putative direct target genes such as *RUNX2*. In a preclinical model, novel_miRNA_2 mimic reduces osteophyte burden and cartilage degradation when administered beginning in early, but not later stages of knee OA, suggesting it has a preventative effect but not a restorative effect. The decline in novel_miRNA_2 levels from early-to late-stage disease may be regulated in part by Hh signaling, which becomes activated in later stages of knee OA and upregulates *RUNX2*, potentially by inhibiting novel_miRNA_2. To our knowledge, this study is the first comprehensive characterization of a circulating novel microRNA in OA.

Reliable biomarkers for early-stage knee OA can transform clinical care by identifying patients for first-line interventions before irreversible joint damage occurs. While serum biomarkers such as cartilage oligomeric matrix protein^32^ and matrix metallopeptidase 3^33^ have been explored in early-stage knee OA, these are not yet in clinical use and are subject to challenges associated with protein biomarkers^34^. MicroRNA biomarkers have been explored in knee OA^35^ and other musculoskeletal diseases^36^, but novel microRNAs offer unique advantages. Novel microRNAs may confer greater specificity and sensitivity as biomarkers since they are considered to be less evolutionarily conserved than established microRNAs, often displaying species-specific or lineage-restricted expression patterns^12,31^. Here, we show circulating novel_miRNA_2 in early-stage knee OA can distinguish both the presence of OA and stage of OA^37^, suggesting it has potential as a diagnostic biomarker. Though not explored in the current study, novel_miRNA_2 may also have potential as a prognostic biomarker^37^, where patients with higher levels of novel_miRNA_2 or longer exposure to novel_miRNA_2 experience slower rates of OA progression. This biomarker potential, together with its protective effects on knee OA, position novel_miRNA_2 as a promising candidate for precision medicine linking patient stratification to targeted interventions, where endogenous novel_miRNA_2 can be used to detect early-stage knee OA and exogenous supplementation can mitigate disease progression.

Our data show novel_miRNA_2 functions as a canonical microRNA in human knee OA subchondral bone and reduces osteophyte burden in mice with knee OA, suggesting it regulates bone-related processes. Structural changes in bone are commonly observed in OA^38^ and can have negative effects on surrounding tissues^39^, including cartilage^40^. As such, preventing abnormal bone-related processes in early-stage knee OA via novel_miRNA_2 offers a viable therapeutic strategy. It remains unknown if novel_miRNA_2 reduces cartilage damage indirectly, via changes in bone, or more directly, via regulation of cartilage-related target genes, but available evidence suggests the former. In early-stage knee OA cartilage, the trends towards elevated novel_miRNA_2 biogenesis we observed were not significant. Consistent with this, a study profiling novel microRNAs in human osteoarthritic chondrocytes did not report the novel_miRNA_2 sequence^41^, though this may be due to the study design. Meanwhile in early-stage knee OA fat pad, we show novel_miRNA_2 biogenesis is elevated. The fat-related target genes we predicted did not show significant differences with novel_miRNA_2 modulation in subchondral bone, but they may be regulated by novel_miRNA_2 in fat pad, as microRNAs are known to have tissue-specific effects^42^. In support of tissue specificity, novel_miRNA_2 had no effect on synovitis in mice with knee OA, and therefore is unlikely to act on synovium. In this study we prioritized bone-related genes previously implicated in OA, yet we predicted nearly 3,000 direct targets of novel_miRNA_2. Thus, there is a need to elucidate the network of novel_miRNA_2 target genes within and across joint tissues to understand its role in OA, including in regulating entirely new disease mechanisms.

The biomarker and therapeutic potential exhibited by novel_miRNA_2 specifically in early-stage knee OA led us to investigate factors governing this stage-dependent activity. Given our focus on subchondral bone and the presence of GLI consensus binding motifs upstream of novel_miRNA_2, we explored Hh signaling, which is known to upregulate *RUNX2* and exacerbate OA^25^. We hypothesize the lack of Hh signal in early-stage knee OA allows novel_miRNA_2 biogenesis to proceed and inhibit *RUNX2*, while the presence of Hh signal in late-stage knee OA reduces novel_miRNA_2 via GLI repressor isoforms^43^, leaving target genes such as *RUNX2* unchecked and promoting aberrant bone-related processes. Under this hypothesis, sustaining novel_miRNA_2 levels from early-to late-stage knee OA is a strategy to mitigate the effects of Hh signaling. This approach may be preferred over inhibition of Hh signaling, which can have off-target effects^44^. Further research is needed to define parameters such as the optimal treatment window for use of novel_miRNA_2 as a therapeutic in knee OA.

To note limitations of this study, validation in larger, independent cohorts is required to assess the generalizability and clinical utility of novel_miRNA_2 as a diagnostic or prognostic biomarker. Additionally, while *in silico* analyses were used to predict direct target genes of novel_miRNA_2, transcriptional regulation must be verified through reporter construct assays, and indirect target genes must be considered. Finally, we do not perform intra-articular delivery of novel_miRNA_2 mimic to target the local joint environment in mice. However, we show intravenous delivery of mimic leads to localization in mouse knee OA tissues, which can be considered a less invasive treatment approach that is amenable to future clinical translation^45^.

In conclusion, this study demonstrates the potential of novel microRNAs to serve as biomarkers and therapeutic targets in complex diseases. We show novel_miRNA_2 is associated with early-stage knee OA, functioning as a canonical microRNA in subchondral bone to regulate bone-related processes. With further development, this microRNA could support precision medicine approaches in OA care, being used as a biomarker to identify early-stage knee OA patients and a therapeutic to prevent disease progression by targeting tissue-specific mechanisms.

## Methods

### Ethical approval and study cohort

Early– and late-stage knee OA biospecimens were obtained from the HFH OA Cohort. Assessed by an orthopedic surgeon, the early-stage knee OA group comprised symptomatic individuals undergoing knee arthroscopy with KL grade 0 or 1^15,29^, and the late-stage knee OA group comprised symptomatic individuals undergoing total knee arthroplasty with KL grade 3 or 4^14^. Non-OA plasma samples were obtained from the Osteoarthritis Initiative (OAI) “Control Cohort”, comprising individuals without knee symptoms, radiographic knee OA, or risk factors for OA. Further details about the OAI are available at: https://nda.nih.gov/oai^16^. Clinicodemographic variables, including age, self-reported sex, BMI, race, and comorbidities were collected and de-identified for all participants (**Supplemental Table 1**). Ethical approval was obtained from the HFH Institutional Review Board (IRB #13995) for the HFH OA Cohort and via the University of California, San Francisco (FWA approval #00000068; IRB approval #10-00532) for the OAI. All methods were performed following the Declaration of Helsinki guidelines^46^.

Briefly, whole blood samples were centrifuged at 3200 x g for 10 minutes at 4°C to obtain plasma. Early– and late-stage knee OA tissues were collected and processed using our published protocols^47^ and included subchondral bone, infrapatellar fat pad, articular cartilage, and synovium. Tissues were collected intraoperatively and dissected under sterile conditions within four hours of surgery. Biospecimens were either used immediately after processing or flash frozen in liquid nitrogen and stored at –80°C until use.

### Real-time quantitative polymerase chain reaction (RT-qPCR)

For plasma, RNA was isolated using the miRNeasy Serum/Plasma Advanced Kit (QIAGEN) according to the manufacturer’s instructions. For tissue explants, RNA was extracted following our published protocols^47^. Briefly, this included tissue homogenization in TRIzol reagent (Thermofisher Scientific) followed by two rounds of chloroform extraction, then precipitation with 100% isopropanol. Pellets were washed with ice-cold 75% ethanol, air-dried, and eluted in nuclease-free water. RNA quality and concentration were determined using a NanoDrop 2000 spectrophotometer (Thermofisher Scientific).

For microRNA quantification, reverse transcription was performed using the TaqMan microRNA Reverse Transcription Kit (Applied Biosystems) with custom TaqMan Small RNA assays (Applied Biosystems). For gene expression analyses, cDNA was synthesized using the High-Capacity cDNA Reverse Transcription Kit (Applied Biosystems). RT-qPCR was performed using a QuantStudio 7 Pro system (Applied Biosystems) with TaqMan Small RNA/Gene Expression Assays (Applied Biosystems) or SYBR Green-based DNA oligonucleotides (Sigma-Aldrich). A list of probes and primers is provided in **Supplemental Table 2**. MiR-24-3p and *GAPDH* were used as internal reference controls for microRNA and gene analyses, respectively, as done previously^8^. Relative expression levels were calculated using the delta-delta-Ct method^48^.

### Novel_miRNA_2 mimic transfection

A custom miRIDIAN microRNA mimic (Dharmacon) corresponding to the novel_miRNA_2 sequence (UCCCUGUUCGGGCGCCACU) was designed for use both *in vitro* and *in vivo*. This synthetic double-stranded RNA oligonucleotide comprised the mature novel_miRNA_2 sequence and its complement, with chemical enhancements to preferentially select the mature novel_miRNA_2 sequence for loading into RISCs. Mimic specificity was confirmed by RT-qPCR standard curve analysis. Primary human subchondral bone explants (∼100 mg/well) were transfected in 24-well plates with 100 nM novel_miRNA_2 mimic or miRIDIAN Human Negative Control Mimic (“negative control”). Transfection was performed using DharmaFECT-1 (Dharmacon) transfection reagent in 500 µL/well serum-free Dulbecco’s Modified Eagle’s Medium (DMEM) supplemented with 1% penicillin-streptomycin (pen-strep, Gibco). To induce Hh pathway activity with an agonist, explants were treated with 10 µM purmorphamine^28^ (APExBio) or dimethyl sulfoxide (DMSO) vehicle for 24 hours, with a subset concurrently transfected with 100 nM novel_miRNA_2 mimic.

### Bioinformatics

Novel_miRNA_2 target genes were predicted by complementary alignment of its seed-sequence (nucleotide position 2-8) to the 3’ UTRs of all human transcripts annotated in Ensembl BioMart (GRCh38). Putative 8mer, 7mer-m8, and 7mer-A1 matches were retained for further analysis as these binding site configurations are recognized as the strongest and most efficient for microRNA-mediated gene repression^49^. Genes were further assessed for microRNA accessibility by computing the minimum free energy of the 70-nucleotide regions upstream and downstream of the seed-sequence binding sites using RNAfold (ViennaRNA package, v.2.5.1)^50^. The mean flanking region delta Gibbs free energy (ΔG) was compared to the average ΔG of all other 70-nucleotide windows within the same 3’ UTR; binding sites with mean flanking region ΔG above the global 3’ UTR average were prioritized^19^. To select target genes most relevant to OA subchondral bone, genes were cross-referenced with the OsteoDIP database (https://ophid.utoronto.ca/OsteoDIP)^20^, filtering for studies conducted in “subchondral bone”, “femoral condyles”, “medial and lateral tibial plateaus”, and “proximal femur”. Pathway core analysis was performed using Ingenuity Pathway Analysis software (QIAGEN)^21^. Putative transcription factor binding sites within the promoter region of *ZNF773* (transcript NM_001304334.2) were identified using the JASPAR (https://jaspar.elixir.no/)^51^ transcription factor motif database.

### AGO2-RNA immunoprecipitation

AGO2-RNA immunoprecipitation experiments were adapted from an established protocol^52^. Briefly, 100 mg primary human knee OA subchondral bone was lysed in NP40 buffer with protease and RNase inhibitors. After pre-clearing, 200 µL lysate was incubated with 3 µg of anti-AGO2 or IgG antibodies bound to pre-blocked Dynabeads™ Protein G (Thermofisher Scientific). IgG was used as a control for the detection of non-specific protein binding. AGO2/IgG complexes were washed and RNA was extracted using our published protocols^47^. RT-qPCR analysis was performed to measure novel_miRNA_2 and U6 small nuclear RNA levels in AGO2– and IgG-bound fractions. U6 small nuclear RNA was used as a non-microRNA control that does not associate with RISCs and therefore should not co-precipitate with AGO2^52^.

### Cell mineralization assay

Murine MC3T3-E1 pre-osteoblasts were seeded in 12-well plates at 6.0 × 10⁴ cells/mL and cultured at 37°C (with 5% CO₂) in Minimum Essential Medium Alpha (α-MEM) supplemented with 10% fetal bovine serum and 1% pen-strep. At 80% confluence, cells were supplemented with 10 mM β-glycerophosphate and 50 µg/mL ascorbic acid-2-phosphate to induce differentiation. Simultaneously, the cells were treated with 20 nM novel_miRNA_2 mimic or negative control. After 21 days of culture, cells were fixed in 10% neutral buffered formalin (NBF) and mineralization was assessed by von Kossa^53^ and Alizarin Red S (ARS) staining^54^ using established protocols. Briefly, for von Kossa, fixed cells were stained with 1% aqueous silver nitrate under ultraviolet light, and unreacted silver was neutralized with 5% sodium thiosulfate. For ARS, fixed cells were stained with 2% ARS (Sigma-Aldrich), then washed thoroughly. Images were captured at 2x magnification using an Eclipse Ti microscope (Nikon). Bound ARS dye was extracted using 10% acetic acid and average absorbance across technical replicates measured at 405 nm using a plate reader (Perkin-Elmer).

### Animal experiments

Male C57BL/6J mice at 11 weeks of age were purchased from Jackson Laboratories and acclimatized for one week prior to surgery. All animals were housed under a 12-hour light/dark cycle at 21°C and 30% humidity. Animals were handled by trained personnel and closely monitored for signs of distress for the duration of the experiments to minimize suffering. Knee OA was induced at 12 week of age using the partial medial meniscectomy (PMX) model as previously described^8^, with a concurrent group of mice undergoing sham surgery. Animals were randomized into treatment groups comprising tail vein injections of 5 µg (in 50 µL 1X PBS) novel_miRNA_2 mimic or negative control delivered weekly according to the regimens shown in **Figure 4A**. At endpoint (8 weeks post-surgery), animals were euthanized via CO_2_ asphyxiation with cervical dislocation secondary assurance of death, and hind limbs were harvested. Animal experiments were approved by the HFH Institutional Animal Care and Use Committee (IACUC #1616).

### Histological analyses

Mouse knee joints were fixed in 10% NBF at room temperature for 4 days followed by decalcification using a 20% ethylenediaminetetraacetic acid (EDTA) solution for 21 days. Samples were paraffin-embedded, coronally sectioned at 5-micron thickness onto charged slides, then deparaffinized and stained with 0.1% Safranin-O. Cartilage damage was assessed using OARSI scoring^55^. Joints were divided into medial and lateral tibial/femoral quadrants and graded from 0 (no cartilage damage) to 6 (>75% cartilage thickness loss) by two independent blinded observers, with the mean maximal quadrant score assigned to each sample. Synovitis was assessed using the Krenn scoring system^56^. Values from 0 (normal) to 3 (severe) were assigned for each of synovial lining thickness, stromal cell density, and inflammatory infiltration, then summed for a maximum score of 9. For osteophytes, cross-sectional areas were manually drawn and areas for the medial tibia and medial femur were quantified and averaged^57^.

To assess novel_miRNA_2 mimic localization via *in situ* hybridization, mouse knee joints were fixed and decalcified as described above. Tissues were then sectioned onto charged RNAse-free slides at 2-micron thickness. Signal was visualized using a custom-designed novel_miRNA_2 miRNAScope probe (Advanced Cell Diagnostics) with the RNAScope 2.5 HD Kit, according to the manufacturer’s instructions.

### Statistical analyses

All data analyses were performed using R 4.4.0 statistical software; specific R packages are available upon request. Unless otherwise noted, all datapoints represent biological replicates. For all experiments, normality was assessed using the Shapiro-Wilk test. For comparisons across three or more groups, statistical significance was determined by one-way analysis of variance (ANOVA) followed by Tukey’s HSD post hoc test or the Kruskal-Wallis test followed by Dunn’s post hoc test. For comparisons between two groups, statistical significance was determined by two-sided unpaired Welch’s T-test, paired Student’s T-test, or Mann-Whitney U-test. Pearson correlation was performed between novel_miRNA_2 levels and KL grade. Multiple linear regression analysis was used to assess associations with covariates. AUCs were generated using the pROC v.1.18.5 R package.

## Data Availability

All data produced in the present work are contained in the manuscript.

## Acknowledgements

MB discloses support for this research from the Postdoctoral/Resident Fellowship awarded by the Orland Bethel Family Musculoskeletal Research Center, University of Pittsburgh Department of Orthopaedic Surgery, and Orthopaedic Research Society. We thank participants of the Henry Ford Health OA Cohort for donating biospecimens.

## Author contributions

MB, TGW, and SAA were involved in study conception and design. MB and TGW performed experimental work and data acquisition. TB, JD, and VM contributed to the Henry Ford Health OA Cohort. MB, TGW, and SAA performed data analysis, interpretation, and drafted the manuscript. All authors contributed to manuscript revisions and approved the final version.

## Supplemental Figures

**Supplemental Figure 1.**
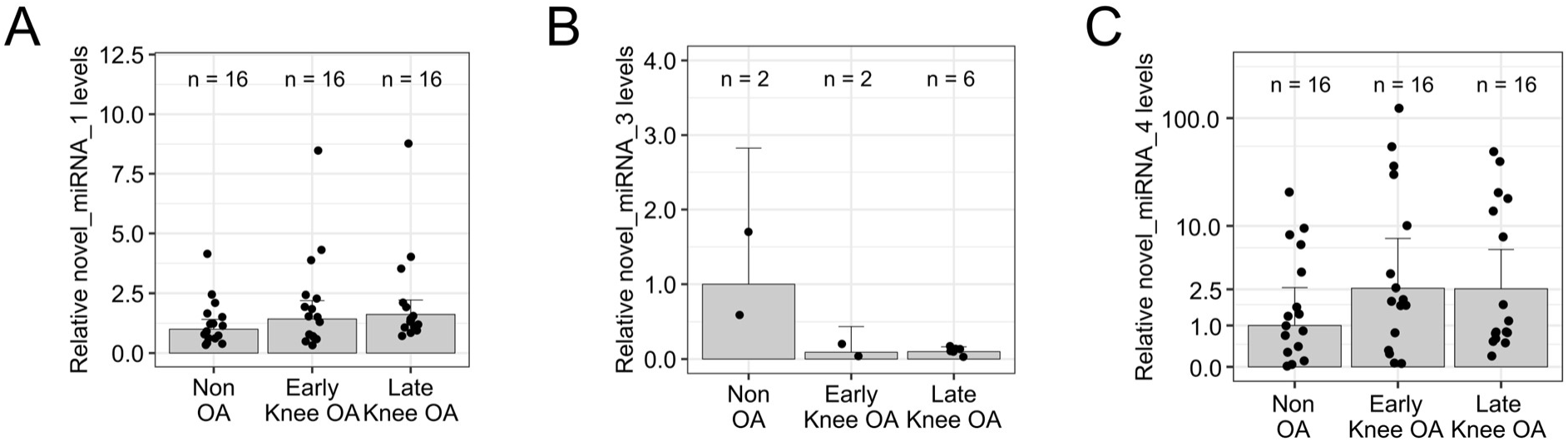
Novel microRNA levels in OA plasma. RT-qPCR for (A) novel_miRNA_1, (B) novel_miRNA_3, and (C) novel_miRNA_4 in the HFH OA Cohort. Data represent the fold-change relative to non-OA controls. Bars represent mean ± 95% confidence interval. Novel_miRNA_3 was not in the detectable range of RT-qPCR for 38/48 of the samples. Significance not detected for any microRNA as assessed by Kruskal-Wallis test, p = 0.232 (novel_miRNA_1), p = N/A (novel_miRNA_3), p = 0.434 (novel_miRNA_4).

**Supplemental Figure 2.**
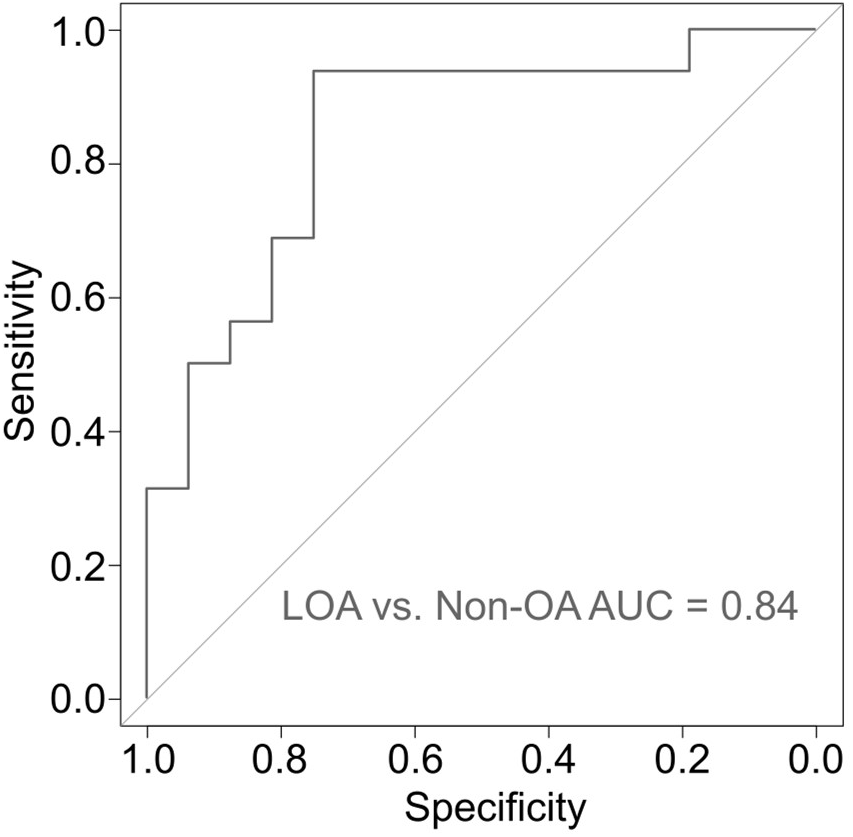
Accuracy of novel_miRNA_2 plasma levels in distinguishing late-stage knee OA (LOA) from non-OA controls. Area under the receiver operating characteristic curve (AUC) analysis.

**Supplemental Figure 3.**
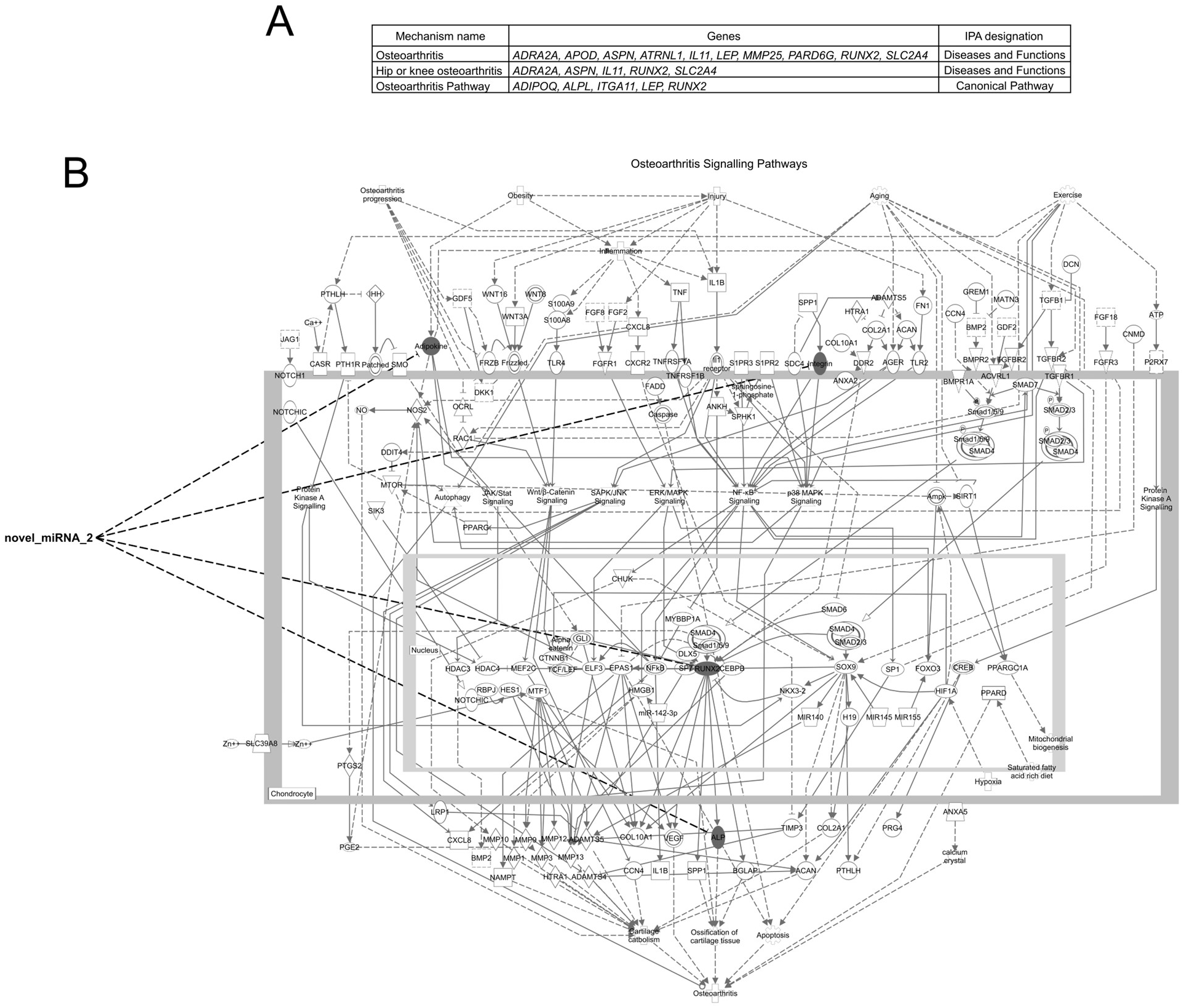
Predicted novel_miRNA_2 target genes in curated ‘Diseases and Functions’ or ‘Canonical Pathways’ related to OA. A) Table of novel_miRNA_2 target genes stratified by Ingenuity Pathway Analysis (IPA) software core analysis results. B) ‘Osteoarthritis Signaling Pathways’ schematic generated by IPA showing predicted novel_miRNA_2 targets as shaded symbols.

**Supplemental Figure 4.**
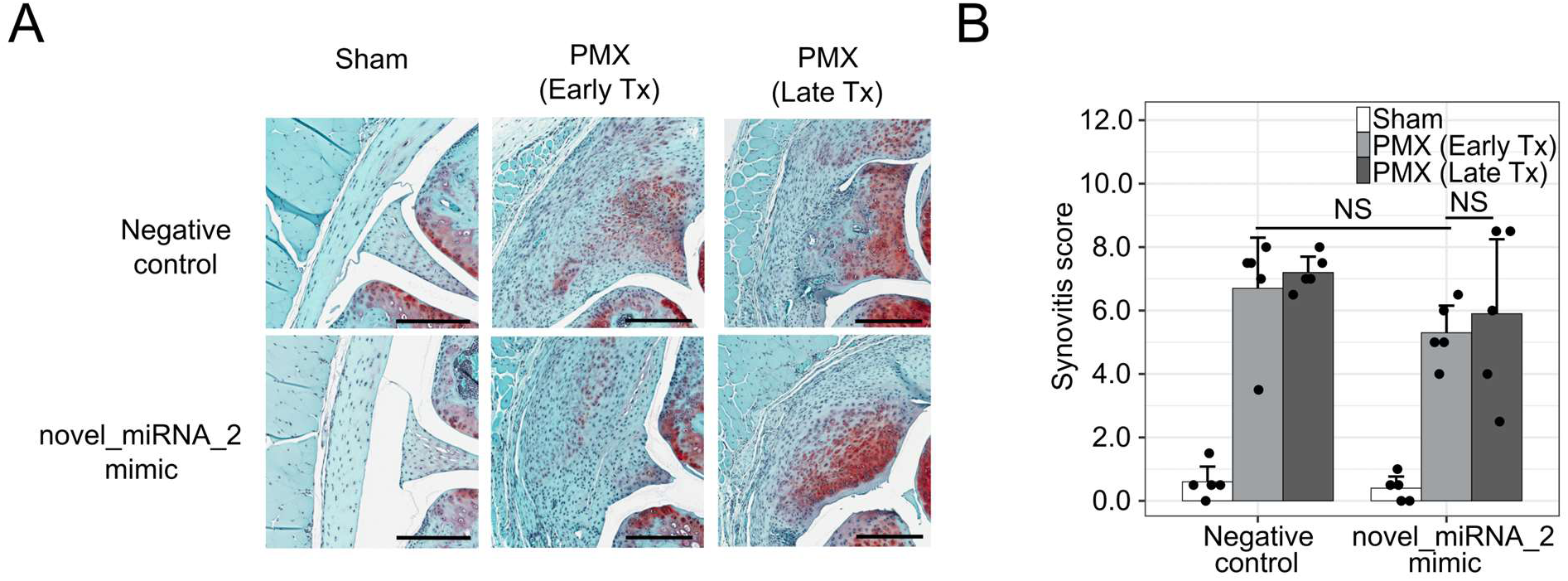
Male mice with knee OA show no change in synovitis following novel_miRNA_2 mimic treatment. A) Representative medial synovium from coronal sections of mouse operated knees. Scale bars = 250 µm. PMX = partial medial meniscectomy. Tx = treatment. B) Krenn synovitis scoring in PMX and sham mice following novel_miRNA_2 mimic or negative control treatments (n = 5). Bars represent mean synovitis score ± 95% confidence interval. NS = not significant as determined by two-sided Mann-Whitney U-test. P-values: p = 0.141 [PMX (Early Tx): Negative control vs. novel_miRNA_2 mimic], p = 0.833 [novel_miRNA_2 mimic: PMX (Early Tx) vs. PMX (Late Tx)].

**Supplemental Table 1.** Clinicodemographic variables of plasma samples used from the HFH OA Cohort. n = sample size, SD = standard deviation, BMI = body mass index. Significance between groups assessed by one-way ANOVA for continuous variables and two-sided Chi-square test for categorical variables. P-values: p = 0.0001 (Age), p = 0.007 (BMI).

|  | Non<br>OA | Early<br>Knee OA | Late<br>Knee OA | p-value |
| --- | --- | --- | --- | --- |
| n (48 total) | 16 | 16 | 16 | - |
| Age (years)<br>Mean (SD) | 58.6<br>(11.2) | 46.6<br>(12.2) | 64.1<br>(8.3) | <0.01 |
| Sex (female)<br>n (%) | 8<br>(50.0) | 7<br>(43.8) | 8<br>(50.0) | 0.92 |
| BMI (kg/m<sup>2</sup>)<br>Mean (SD) | 27.0<br>(3.2) | 27.6<br>(5.1) | 31.9<br>(5.2) | <0.01 |
| Race (white)<br>n (%) | 15<br>(93.8) | 10<br>(62.5) | 13<br>(81.2) | 0.24 |

**Supplemental Table 2.**
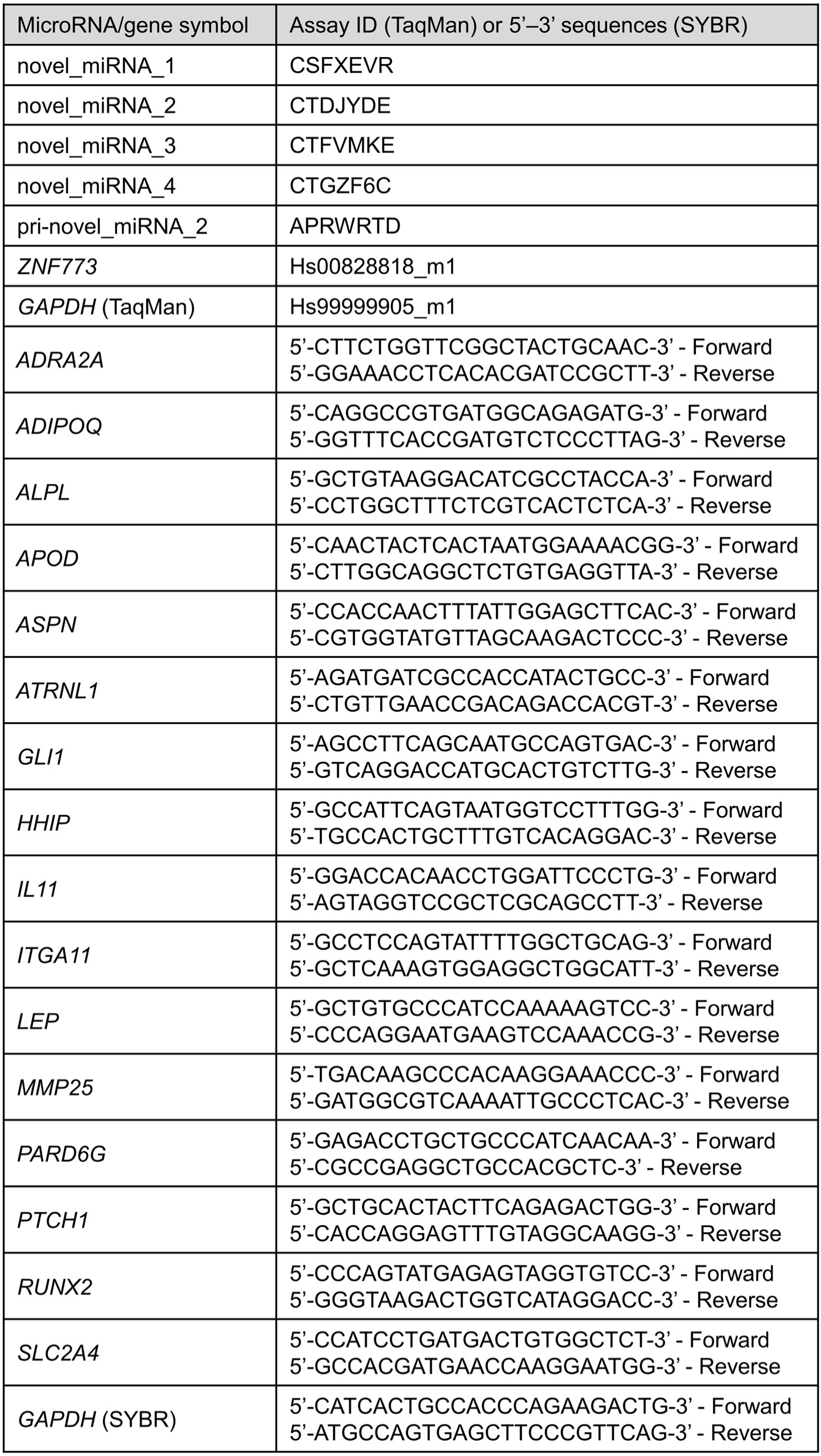
List of probes and primers used for RT-qPCR experiments.

